# Clinical Evaluation of the Novel Rapid Nucleic Acid Amplification Point-of-Care Test (Smart Gene SARS-CoV-2) in the analysis of Nasopharyngeal and Anterior Nasal samples

**DOI:** 10.1101/2021.08.25.21262583

**Authors:** Yoshihiko Kiyasu, Masato Owaku, Yusaku Akashi, Yuto Takeuchi, Kenji Narahara, Sunao Mori, Takashi Nagano, Shigeyuki Notake, Atsuo Ueda, Koji Nakamura, Hiroichi Ishikawa, Hiromichi Suzuki

**Affiliations:** Division of Infectious Diseases, Department of Medicine, Tsukuba Medical Center Hospital, 1-3-1 Amakubo Tsukuba, Ibaraki 305-8558, Japan; Department of Infectious Diseases, University of Tsukuba Hospital, 2-1-1 Amakubo, Tsukuba, Ibaraki 305-8576, Japan; Mizuho Medy Co., Ltd., 5-4 Fujinoki-machi, Tosu City, Saga 841-0048, Japan; Akashi Internal Medicine Clinic, 3-1-63 Asahigaoka, Kashiwara, Osaka 582-0026, Japan; Department of Clinical Laboratory, Tsukuba Medical Center Hospital, 1-3-1 Amakubo, Tsukuba, Ibaraki 305-8558, Japan; Department of Respiratory Medicine, Tsukuba Medical Center Hospital, 1-3-1 Amakubo Tsukuba, Ibaraki 305-8558, Japan; Department of Infectious Diseases, Faculty of Medicine, University of Tsukuba, 1-1-1 Tennodai, Tsukuba, Ibaraki 305-8575, Japan

**Keywords:** point-of-care test, Smart Gene, nucleic acid amplification test, QProbe, SARS-CoV-2, COVID-19

## Abstract

**Introduction:** Smart Gene is a point-of-care (POC)-type automated molecular testing platform that can be performed with 1 minute of hands-on-time. Smart Gene SARS-CoV-2 is a newly developed Smart Gene molecular assay for the detection of SARS-CoV-2. The analytical and clinical performance of Smart Gene SARS-CoV-2 has not been evaluated.

**Methods:** Nasopharyngeal and anterior nasal samples were prospectively collected from subjects referred to the local PCR center from March 25 to July 5, 2021. Two swabs were simultaneously obtained for the Smart Gene SARS-CoV-2 assay and the reference real-time RT-PCR assay, and the results of Smart Gene SARS-CoV-2 were compared to the reference real-time RT-PCR assay.

**Results:** Among a total of 1150 samples, 68 of 791 nasopharyngeal samples and 51 of 359 anterior nasal samples were positive for SARS-CoV-2 in the reference real-time RT-PCR assay. In the testing of nasopharyngeal samples, Smart Gene SARS-CoV-2 showed the total, positive and negative concordance of 99.2% (95% confidence interval [CI]: 98.4–99.7%), 94.1% (95% CI: 85.6–98.4%) and 99.7% (95% CI: 99.0–100%), respectively. For anterior nasal samples, Smart Gene SARS-CoV-2 showed the total, positive and negative concordance of 98.9% (95% CI: 97.2–99.7%), 98.0% (95% CI: 89.6–100%) and 99.0% (95% CI: 97.2–99.8%), respectively. In total, 5 samples were positive in the reference real-time RT-PCR and negative in Smart Gene SARS-CoV-2, whereas 5 samples were negative in the reference real-time RT-PCR and positive in Smart Gene SARS-CoV-2.

**Conclusion:** Smart Gene SARS-CoV-2 showed sufficient analytical performance for the detection of SARS-CoV-2 in nasopharyngeal and anterior nasal samples.

## Introduction

Since its emergence in late 2019, severe acute respiratory syndrome coronavirus 2 (SARS-CoV-2) has spread worldwide [1]. A quick and accurate diagnosis is essential for the clinical management of patients and infection control measures [2]. The expanding pandemic has increased the demand for laboratory tests and large-scale testing. Facility-based platforms using a high-capacity automated testing system are advantageous for meeting this demand. However, the time to perform the test and transport the samples to the laboratory are barriers to obtaining results in a timely manner.

A point-of-care (POC) test, which does not require extensive equipment or skilled technicians and which provides results in a short period, is beneficial for overcoming the disadvantages of large-scale laboratory testing. POC tests are helpful in situations like outbreaks in long-term care facilities, where an immediate diagnosis is needed [2]. Antigen tests are quicker and easier to use than nucleic acid amplification tests (NAAT) and are widely used as POC tests. However, a recent systematic meta-analysis of antigen test performance in real-world settings revealed that sensitivity ranges from 28.9% to 98.3% according to demographic features, viral load, and symptom state [3]. Thus, rapid and more reliable diagnostic tools are required.

Smart Gene SARS-CoV-2 (Mizuho Medy Co., Ltd., Tosu City, Saga, Japan) is a novel test kit for Smart Gene, an automated molecular testing platform that utilizes the quenching probe (QProbe) method. Because of its small size and fast turnaround time, Smart Gene can be installed in clinics and used for POC testing platforms, and has been used to diagnose pediatric respiratory infection of *Mycoplasma pneumoniae* [4]. In this prospective study, we evaluate the clinical performance of Smart Gene SARS-CoV-2 for nasopharyngeal and anterior nasal samples collected at a local PCR center.

## Patients and Methods

In this study, we enrolled participants and collected samples at the PCR center in Tsukuba Medical Center Hospital (TMCH) between March 25 and July 5, 2021, which intensively collected nasopharyngeal samples for the PCR analysis of SARS-CoV-2 [5, 6, 7, 8, 9]. Subjects referred by 59 primary care facilities and a local public health center, as well as TMCH healthcare workers with suspected SARS-CoV-2 infection based on symptoms or a known contact history with COVID-19 confirmed/suspected patients were prospectively enrolled.

For the evaluation of nasopharyngeal samples, we obtained one additional nasopharyngeal sample for the evaluation of Smart Gene SARS-CoV-2, as previously described [6, 7]. For the evaluation of anterior nasal samples, we obtained two additional anterior nasal samples, as previously described [5], for the Smart Gene SARS-CoV-2 and reference real-time RT-PCR assays. The Sponge swab^™^(NIPRO, Osaka, Japan), equipped with a Smart Gene SARS-CoV-2 kit, was used for obtaining samples for the Smart Gene SARS-CoV-2 assay and the FLOQ swab^™^ (Copan Italia S.p.A., Brescia, Italy) was used for obtaining samples for in-house reverse transcription (RT)-PCR and reference real-time RT-PCR assays.

All subjects provided their consent for enrollment. If multiple samples were collected from the same subject, they were all treated as individual samples. The ethics committee of TMCH approved the present study (approval number: 2021-008).

### Smart Gene SARS-CoV-2 assay

The evaluation of Smart Gene SARS-CoV-2 was performed using fresh samples according to the manufacturer’s instructions, as described in the package insert. An overview of the test procedure is shown in Figure 1. Briefly, the collected sample with the sponge swab was suspended with 1 mL of extraction reagent solution in a vial, and the suspended sample was dripped into the cartridge. The cartridge was inserted into the analyzer and the fully automated PCR analysis was performed after a few touch-steps.

**Figure 1.**
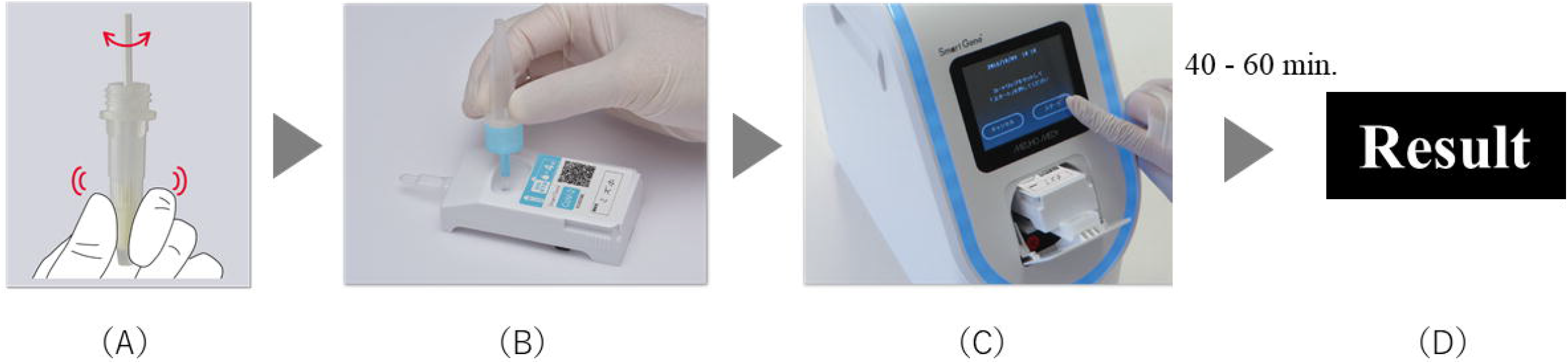
Smart Gene SARS-CoV-2 assay workflow (A) Suspend the collected sample in the extraction reagent solution vial. (B) Drop the suspended sample on the cartridge. (C) Set cartridge into the analyzer and touch the start button on the screen. (D) The analyzer automatically performs nucleic acid extraction and amplification steps, and reports the test result and the Ct value within 40–60 min.

### Reference real-time RT-PCR assay for SARS-CoV-2

The swabs were suspended in 3 mL of Universal Transport Medium^™^ (UTM^™^) (Copan diagnostics, Brescia, Italy). Purification and RNA extraction from UTM^™^ samples were performed with magLEAD^®^ 6gC (Precision System Science, Chiba, Japan). The purified samples were evaluated by GENECUBE^®^ and GENECUBE^®^ HQ SARS-CoV-2 as an in-house RT-PCR assay [8]. The purified samples were frozen at -80 °C and transferred to Mizuho Medy for the reference real-time RT-PCR assay. The N2 primer/probe set (Nihon Gene Research Laboratories, Miyagi, Japan) was employed for the reference real-time RT-PCR assay, as suggested by the “Manual for the Detection of Pathogen 2019-nCoV Ver. 2.9.1” issued by the National Institute of Infectious Diseases of Japan (NIID) [10]. The reference real-time RT-PCR assays were performed on a LightCycler^®^ Nano System (Roche Diagnostics, Rotkreuz, Switzerland) using One Step PrimeScript^™^ III RT-qPCR Mix (Takara Bio, Kusatsu, Japan) with the following cycling conditions: reverse transcription at 52°C for 5 min and 95°C for 10 s, and 45 cycles at 95°C for 5 s and at 60°C for the 30s. The absolute viral copy number was determined by serially diluted RNA control targeting the N2 gene of SARS-CoV-2 (Nihon Gene Research Laboratories).

### Validation of discordant cases

For discordant samples that were negative by Smart Gene SARS-CoV-2 and positive by the reference real-time RT-PCR assay, the stored UTM samples were re-evaluated by Smart Gene SARS-CoV-2. Three hundred microliters of each UTM sample were added to the extract reagent solution, and subsequent operations were performed according to the manufacturer’s instructions. These measurements were performed in duplicate. For discordant samples that were positive by Smart Gene SARS-CoV-2 and negative by the reference real-time RT-PCR assay, each stored UTM sample was evaluated by GeneXpert^®^ for SARS-CoV-2 (Cepheid, Sunnyvale, CA, USA) according to the manufacturer’s instructions.

### Limits of detection of Smart Gene SARS-CoV-2 and reference real-time RT-PCR

The limits of detection (LOD) of Smart Gene SARS-CoV-2 and reference real-time RT-PCR were evaluated using the AccuPlex SARS-CoV-2 Verification Panel (SeraCare Life Sciences, Inc., Milford, MA, USA), and negative nasopharyngeal swabs.

For sample preparation, negative nasopharyngeal swabs were obtained from 4 healthy volunteers and were suspended into either 3 mL of UTM for the reference real-time RT-PCR assay or 1 mL of extraction reagent solution of Smart Gene SARS-CoV-2. Each suspended sample was pooled, and a total of 2 pooled samples for each solution were prepared. Then, each pooled solution was divided into 4 groups by adding diluted AccuPlex SARS-CoV-2 Verification Panel (Supplementary Figure 1), and negative samples were also prepared. The viral concentration for UTM was 1,824 copies/mL, 912 copies/ mL, 456 copies/mL, 228 copies/ mL, and 0 copy/mL respectively.

For evaluation of the LOD, a total of 24 replicate analyses (8 for pooled sample 1, 8 for pooled sample 2, 8 for UTM) for the reference real-time RT-PCR assay, and 12 replicate analyses (4 for pooled sample 1, 4 for pooled sample 2, 4 for extraction reagent solution) for Smart Gene SARS-CoV-2 were performed for each viral concentration. The N2 primer/probe set of the NIID method was used for the reference real-time RT-PCR assay. The lowest viral load at which a 100% detection rate was achieved was considered to be the LOD of the assay.

### Statistical analyses

The 95% confidence intervals (CIs) of total, positive and negative concordance rate between Smart Gene SARS-CoV-2 and reference real-time RT-PCR were calculated using the Clopper and Pearson method. The correlation of Ct values between the reference real-time RT-PCR assay and Smart Gene SARS-CoV-2 was evaluated using Pearson’s correlation coefficient. All statistical analyses were conducted using the R 4.0.3 software program (www.r-project.org).

## Results

During the study period, 1150 samples were collected, of which 791 were nasopharyngeal samples and 359 were anterior nasal samples (Table 1a,b,c). Sixty-eight of 791 (8.6 %) nasopharyngeal samples and 51 of 359 (14.2 %) anterior nasal samples were positive for SARS-CoV-2 by the reference real-time RT-PCR assay. All of the results of the in-house RT-PCR assays were in concordance with the results of the reference real-time RT-PCR assays.

**Table 1a.** The concordance rate between Smart Gene SARS-CoV-2 and reference real-time RT-PCR using nasopharyngeal samples and anterior nasal samples

|  |  | Reference real-time RT-PCR |  |
| --- | --- | --- | --- |
|  |  | Positive | Negative |
| Smart Gene SARS-CoV-2 | Positive | 114 | 5 |
|  | Negative | 5 | 1026 |
| Positive concordance rate (%) |  | 95.8 ( 90.5 - 98.6 ) |  |
| Negative concordance rate (%) |  | 99.5 ( 98.9 - 99.8 ) |  |
| Total concordance rate (%) |  | 99.1 ( 98.4 - 99.6 ) |  |

Data in parentheses are 95% confidence intervals.
*RT-PCR* reverse transcription polymerase chain reaction, *SARS-CoV-2* severe acute respiratory syndrome coronavirus 2.

**Table 1b.**
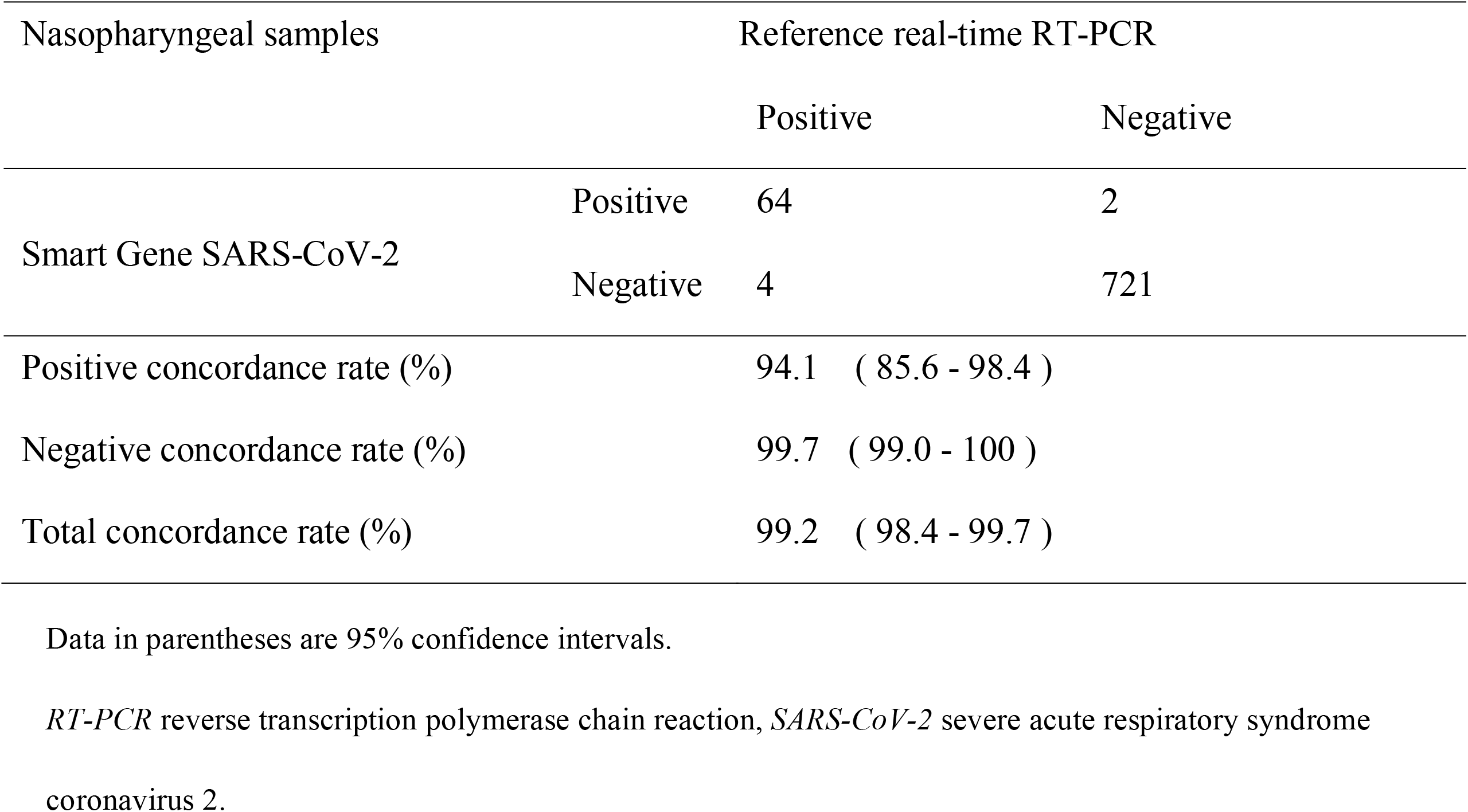
The concordance rate between Smart Gene SARS-CoV-2 and reference real-time RT-PCR using nasopharyngeal samples

| Nasopharyngeal samples |  | Reference real-time RT-PCR |  |
| --- | --- | --- | --- |
|  |  | Positive | Negative |
| Smart Gene SARS-CoV-2 | Positive | 64 | 2 |
|  | Negative | 4 | 721 |
| Positive concordance rate (%) |  | 94.1 ( 85.6 - 98.4 ) |  |
| Negative concordance rate (%) |  | 99.7 ( 99.0 - 100 ) |  |
| Total concordance rate (%) |  | 99.2 ( 98.4 - 99.7 ) |  |

Data in parentheses are 95% confidence intervals.
*RT-PCR* reverse transcription polymerase chain reaction, *SARS-CoV-2* severe acute respiratory syndrome coronavirus 2.

**Table 1c.** The concordance rate between Smart Gene SARS-CoV-2 and reference real-time RT-PCR using anterior nasal samples

| Anterior nasal samples |  | Reference real-time RT-PCR |  |
| --- | --- | --- | --- |
|  |  | Positive | Negative |
| Smart Gene SARS-CoV-2 | Positive | 50 | 3 |
|  | Negative | 1 | 305 |
| Positive concordance rate (%) |  | 98.0 ( 89.6 - 100 ) |  |
| Negative concordance rate (%) |  | 99.0 ( 97.2 - 99.8 ) |  |
| Total concordance rate (%) |  | 98.9 ( 97.2 – 99.7 ) |  |

Data in parentheses are 95% confidence intervals.
*RT-PCR* reverse transcription polymerase chain reaction, *SARS-CoV-2* severe acute respiratory syndrome coronavirus 2.

### Results of the limit of detection tests using recombinant SARS-CoV-2

The results of the LOD tests using recombinant SARS-CoV-2 are shown in Table 2a and b. For the current LOD evaluation, the reference real-time RT-PCR LOD showed positive results in all spiked samples down to 912 copies/mL. Smart Gene SARS-CoV-2 provided positive results in all samples with viral concentration corresponding to 456 copies/mL in UTM samples for the reference real-time RT-PCR assay.

**Table 2a.** Detailed results of the LOD test for the Smart Gene SARS-CoV-2 and reference real-time RT-PCR assays

| Viral loads of<br>UTM samples<br>(Copies / mL) | Sample | Reference real-time RT-PCR<sup>a</sup><br>(NIID method, N2 gene) |  | Smart Gene SARS-CoV-2<sup>a</sup> |  |
| --- | --- | --- | --- | --- | --- |
|  |  | positive / test | Average Ct value | positive / test | Average Ct value |
|  |  | (Detection rate) |  | (Detection rate) |  |
| 1824 | Total | 24/24 (100) | - | 12/12 (100) | - |
|  | UTM or extraction reagent solution | 8/8 (100) | 33.6 | 4/4 (100) | 39.5 |
|  | Pooled nasopharyngeal sample 1 | 8/8 (100) | 33.6 | 4/4 (100) | 39.5 |
|  | Pooled nasopharyngeal sample 2 | 8/8 (100) | 33.9 | 4/4 (100) | 39.5 |
| 912 | Total | 24/24 (100) | - | 12/12 (100) | - |
|  | UTM or extraction reagent solution | 8/8 (100) | 34.5 | 4/4 (100) | 40.5 |
|  | Pooled nasopharyngeal sample 1 | 8/8 (100) | 35.0 | 4/4 (100) | 40.0 |
|  | Pooled nasopharyngeal sample 2 | 8/8 (100) | 34.8 | 4/4 (100) | 40.8 |
| 456 | Total | 22/24 (91.7) | - | 12/12 (100) | - |
|  | UTM or extraction reagent solution | 8/8 (100) | 35.8 | 4/4 (100) | 41.3 |
|  | Pooled nasopharyngeal sample 1 | 7/8 (87.5) | 35.7 | 4/4 (100) | 40.5 |
|  | Pooled nasopharyngeal sample 2 | 7/8 (87.5) | 35.6 | 4/4 (100) | 40.3 |
| 228 | Total | 21/24 (87.5) | - | 7/12 (58.3) | - |
|  | UTM or extraction reagent solution | 7/8 (87.5) | 36.1 | 3/4 (75.0) | 43.3 |
|  | Pooled nasopharyngeal sample 1 | 7/8 (87.5) | 36.5 | 2/4 (50.0) | 41.0 |
|  | Pooled nasopharyngeal sample 2 | 7/8 (87.5) | 35.6 | 2/4 (50.0) | 42.0 |

| 0 | Total | 0/24 (0) | - | 0/12 (0) | - |
| --- | --- | --- | --- | --- | --- |
|  | UTM or extraction reagent solution | 0/8 (0) | ND | 0/4 (0) | ND |
|  | Pooled nasopharyngeal sample 1 | 0/8 (0) | ND | 0/4 (0) | ND |
|  | Pooled nasopharyngeal sample 2 | 0/8 (0) | ND | 0/4 (0) | ND |

*Ct* cycle threshold, *LOD* limit of detection, *ND* not detected, *NIID* National Institute of Infectious Diseases, *RT-PCR* reverse transcription polymerase chain reaction, *UTM* Universal Transport Medium
<sup>a</sup>For the LOD evaluation, a total of 24 replicate analyses (8 for pooled sample 1, 8 for pooled sample 2, 8 for UTM) for the reference real-time RT-PCR assay, and a total of 12 replicate analyses (4 for pooled sample 1, 4 for pooled sample 2, 4 for extraction reagent solution) for the Smart Gene SARS-CoV-2 assay were performed for each viral concentration.

**Table 2b.** Summary of the results of the LOD test for the reference real-time RT-PCR and Smart Gene SARS-CoV-2 assays

| Viral loads of UTM<br>samples (Copies /<br>mL) | Reference real-time RT-PCR<sup>a</sup> | Smart Gene SARS-CoV-2<sup>a</sup> |
| --- | --- | --- |
|  | (NIID method, N2 gene) |  |
|  | positive / test<br>(Detection rate) | positive / test<br>(Detection rate) |
| 1824 | 24/24 (100) | 12/12 (100) |
| 912 | 24/24 (100) | 12/12 (100) |
| 456 | 22/24 (91.7) | 12/12 (100) |
| 228 | 21/24 (87.5) | 7/12 (58.3) |
| 0 | 0/24 (0) | 0/12 (0) |

*ND* not detected, *NIID* National Institute of Infectious Diseases, *RT-PCR* reverse transcription polymerase chain reaction
<sup>a</sup> For the LOD evaluation, a total of 24 replicate analyses (8 for pooled sample 1, 8 for pooled sample 2, 8 for UTM) for the reference real-time RT-PCR assay, and a total of 12 replicate analyses (4 for pooled sample 1, 4 for pooled sample 2, 4 for extraction reagent solution) for the Smart Gene SARS-CoV-2 assay were performed for each viral concentration.

### Performance evaluation of Smart Gene SARS-CoV-2

The results of the performance evaluation of Smart Gene SARS-CoV-2 are shown in 1a (total samples; nasopharyngeal samples and anterior nasal samples), Tables 1b, (nasopharyngeal samples), and 1c (anterior nasal samples).

For nasopharyngeal samples (Table 1b), sixty-eight nasopharyngeal samples were positive by reference real-time RT-PCR and sixty-six were positive by Smart Gene SARS-CoV-2. A total of 64 samples were positive both by reference real-time RT-PCR and Smart Gene SARS-CoV-2. Thus, the total, positive, and negative concordance rate between the two assays were 99.2% (95% CI: 98.4–99.7%), 94.1% (95% CI: 85.6–98.4%), 99.7% (95% CI: 99.0–100%), respectively (Table 1b).

For anterior nasal samples (Table 1c), 51 anterior nasal samples were positive by reference real-time RT-PCR and 53 were positive by Smart Gene SARS-CoV-2. A total of 50 samples were positive both by reference real-time RT-PCR and Smart Gene SARS-CoV-2. Therefore, the total, positive, and negative concordance rate between the two assays were 98.9% (95% CI: 97.2–99.7%), 98.0% (95% CI: 89.6–100%), and 99.0% (95% CI: 97.2–99.8%), respectively (Table 1c).

The overall evaluation of nasopharyngeal and anterior nasal samples is summarized in Table 1a. The total, positive and negative concordance rate between the two assays were 99.1% (95% CI: 98.4–99.6%), 95.8% (95% CI: 90.5–98.6%), and 99.5% (95% CI: 98.9–99.8%), respectively.

### Evaluation of samples for which Smart Gene SARS-CoV-2 and reference real-time RT-PCR showed discordant results

The detailed evaluations of samples for which the Smart Gene SARS-CoV-2 and reference real-time RT-PCR assays showed discordant results are summarized in Table 3a (nasopharyngeal samples) and 3b (anterior nasal samples).

**Table 3a.**
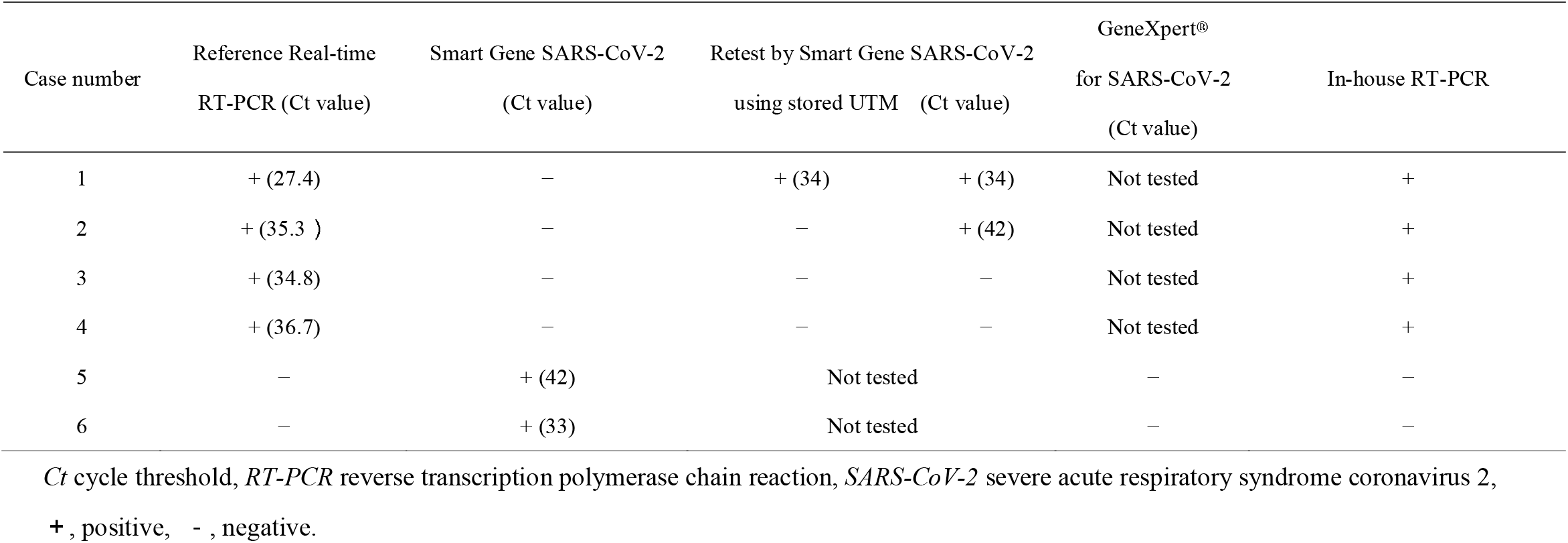
Detailed data of the six cases for which the Smart Gene SARS-CoV-2 and reference real-time RT-PCR assays showed discordant results for nasopharyngeal samples

**Table 3b.**
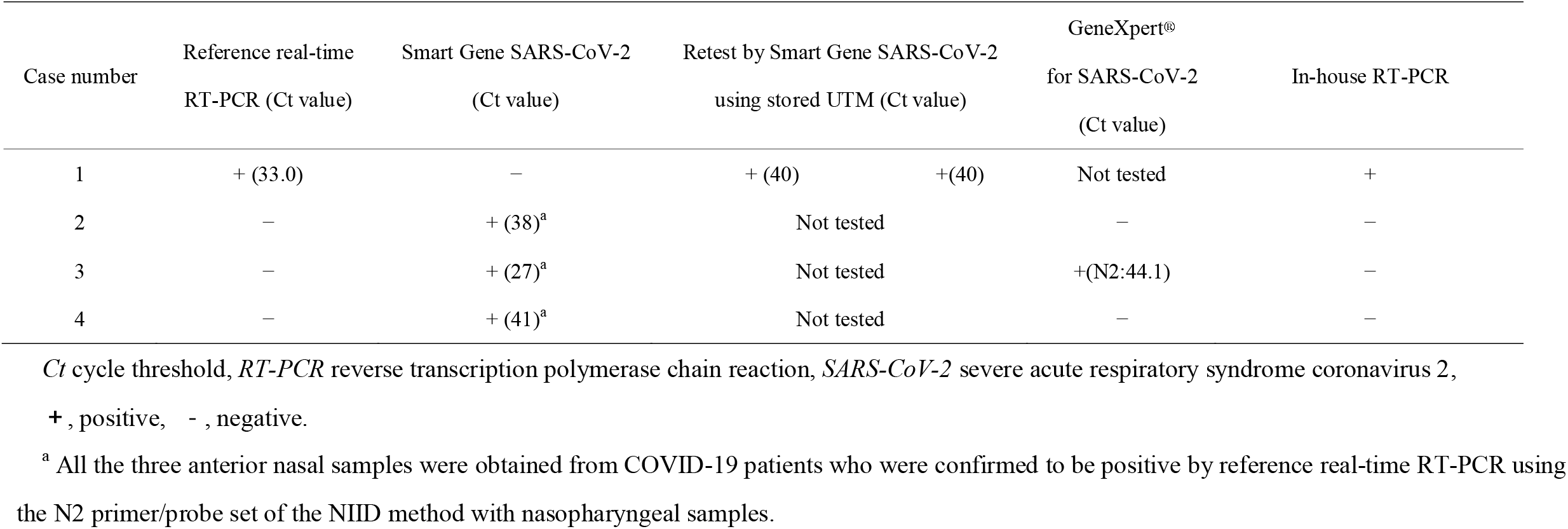
Detailed data of the four cases for which the Smart Gene SARS-CoV-2 and reference real-time RT-PCR assays showed discordant results for anterior nasal samples

For six discordant nasopharyngeal samples, four samples were positive by reference real-time RT-PCR and negative by Smart Gene SARS-CoV-2 and all of the samples were positive by in-house RT-PCR. Of the four samples, two samples were positive by the re-evaluation by Smart Gene SARS-CoV-2 using stored discordant UTM samples. Two samples with negative reference real-time RT-PCR results and positive Smart Gene SARS-CoV-2 results were negative by GeneXpert^®^ using stored UTM samples.

For four anterior nasal samples, one sample was positive by reference real-time RT-PCR and negative by Smart Gene SARS-CoV-2 and the sample were positive by in-house RT-PCR. The sample was positive in a re-evaluation by Smart Gene SARS-CoV-2 using stored discordant UTM samples. For three samples with negative reference real-time RT-PCR results and positive Smart Gene SARS-CoV-2 results, one sample was positive and two samples were negative by GeneXpert^®^ using stored UTM samples. All three anterior nasal samples were obtained from COVID-19 patients confirmed by positive reference real-time RT-PCR using the N2 primer/probe set of the NIID method with nasopharyngeal samples.

### Comparison of the Ct value between Smart Gene SARS-CoV-2 and reference real-time RT-PCR

The comparison of the Ct values of Smart Gene SARS-CoV-2 and the reference real-time RT-PCR assay are plotted in Figure 2. The calculation of Pearson’s correlation coefficient revealed a significant correlation (R=0.81, *p* values < 0.001) between the reference RT-PCR and Smart Gene SARS-CoV-2 assays. The median Ct value was 29.0 (IQR: 25.0-34.8) for Smart Gene SARS-CoV-2 and 23.0 (IQR: 19.6-28.5) for the reference real-time RT-PCR assay. The median difference of Ct values between Smart Gene SARS-CoV-2 and the reference real-time RT-PCR assay was 6.0 (IQR: 3.9-7.8). The Ct value of each sample is listed in Supplementary Table 1 and Supplementary Figure 2.

**Figure 2.**
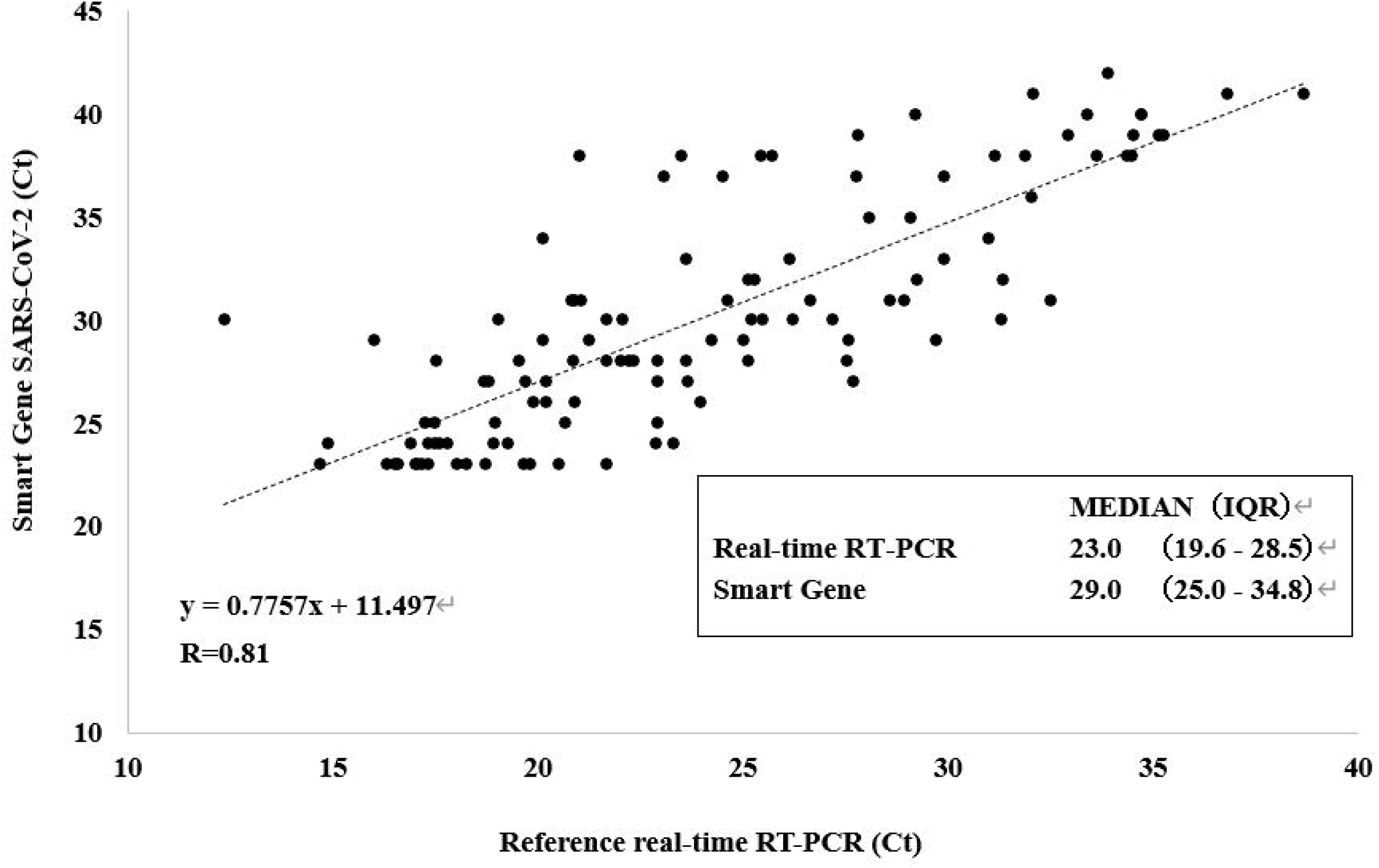
Comparison of the cycle threshold (Ct) values between the Smart Gene SARS-CoV-2 and reference real-time RT-PCR assays. Pearson’s correlation coefficient (R) between two tests was 0.81.

## Discussion

The comparison between the Smart Gene SARS-CoV-2 and reference real-time RT-PCR method of the National Institute of Infectious Diseases (NIID) demonstrated that Smart Gene SARS-CoV-2 has equal analytical performance in the detection of SARS-CoV-2 in fresh nasopharyngeal samples and anterior nasal samples. This was also confirmed through the LOD evaluation. A strong correlation was indicated between the Ct value of Smart Gene SARS-CoV-2 and the reference real-time RT-PCR assay. The median difference of Ct values between the Smart Gene SARS-CoV-2 and reference real-time RT-PCR assays was 6.0.

Through the comparison with 1150 samples, the results of the Smart Gene SARS-CoV-2 and reference real-time RT-PCR assays deviated in 10 samples. For 5 samples with positive reference real-time RT-PCR results and negative Smart Gene SARS-CoV-2 results, GeneXpert^®^ showed positive results; thus, the Smart Gene SARS-CoV-2 assay was considered to have provided false-negative results. For 5 samples with negative reference real-time RT-PCR results and positive Smart Gene SARS-CoV-2 results, 3 of the 5 samples were anterior nasal samples, which were obtained from patients with SARS-CoV-2 infection that was confirmed by a positive nasopharyngeal real-time RT-PCR assay; thus, the reference real-time RT-PCR of anterior nasal samples was considered to have provided false-negative results. Some factors may have caused the inconsistency between the two molecular assays can be explained. The swabs used for the Smart Gene SARS-CoV-2 and the reference real-time RT-PCR assays were obtained separately. Other variables such as the possibility of a non-specific reaction in the Smart Gene SARS-CoV-2 assay, RNA extraction procedures, the interval from sample collection and testing, and randomness in low virus load samples may have influenced the resuls [11].

Currently, several POC-type molecular assays have been available worldwide [3], and their sensitivities and specificities were generally as high as above 90% [3, 12, 13]. The Smart Gene SARS-CoV-2 is characterized by the simplified operation, the short hands-on time of less than 1 minute, and the small equipment size [4]. Besides, our study indicated that Smart Gene SARS-CoV-2 has comparable diagnostic performance to other POC molecular tests [3, 12, 13]. In the evaluations of LOD and clinical performance, Smart Gene SARS-CoV-2 provided almost identical results to the reference real-time RT-PCR.

Another feature of the system is to display Ct values of samples. The appropriate interpretation of Ct values is important in clinical contexts. The Ct values may predict not only the viral loads in samples but also the infectivity or disease severity of patients with SARS-CoV-2 [14]. The current evaluation observed the linear Ct value correlation (*p* values < 0.001) between Smart Gene SARS-CoV-2 and the reference real-time RT-PCR (national standard method in Japan). Nevertheless, the provided Ct values should be carefully read. The Smart Gene SARS-CoV-2 showed 6 points higher Ct values in median than the reference RT-PCR, and marked differences existed in some samples (Supplementary Figure 2 and Supplementary Table 1).

Our study was associated with some limitations. First, we did not analyze the sequences of the viruses detected in this study. Thus, the effect of genetic mutation on the performance of Smart Gene SARS-CoV-2 was not evaluated. Second, this study was conducted in a limited season and area, and it may be necessary to verify whether similar results can be obtained throughout the year or in another area. Finally, the reference real-time RT-PCR assay used frozen stored purified extraction, and such storage and transportation methods may have affected the test results.

In conclusion, current study showed that Smart Gene SARS-CoV-2 had sufficient analytical performance in the detection of SARS-CoV-2 in nasopharyngeal and anterior nasal samples.

## Data Availability

Some restrictions will apply.

## Acknowledgments

We thank Yoko Ueda, Mio Matsumoto, Suwako Kikuchi, Kaoru Kuriiwa, Masaomi Matsubayashi, Yumiko Tanaka, Mika Yaguchi, Shoko Yoshiwara, Asami Sugie, Shiori Kanoya, Kodai Tayama, Kazuya Ishiguro, Toshiki Yoshizawa, Norihiko Terada, Naoki Tanimura and the staff in the Department of Clinical Laboratory of Tsukuba Medical Center Hospital for their intensive support of this study. We thank all of the medical institutions for providing their patients’ clinical information.

## Conflicts of interest

Mizuho Medy provided fees for research expenses and provided Smart Gene SARS-CoV-2 free of charge. Masato Owaku, Kenji Narahara, Sunao Mori, Takashi Nagano are employed by Mizuho Medy, the developer of Smart Gene SARS-CoV-2. Hiromichi Suzuki received a consultation fee from Mizuho Medy.

---

Supplementary Figure 1. Illustration of the sample preparation for the LOD test. Pooled negative nasopharyngeal samples were prepared with nasopharyngeal swabs obtained from healthy volunteers and Universal Transport Medium (UTM) (Copan diagnostics, Brescia, Italy) for the reference real-time RT-PCR assay or extraction reagent solution of the Smart Gene SARS-CoV-2. AccuPlex SARS-CoV-2 Verification Panel (SeraCare Life Sciences, Inc., Milford, MA, USA) was serially diluted 2-fold, yielding dilutions with viral concentrations of 8 × 10^4^ copies/mL, 4 × 10^4^ copies/mL, 2 × 10^4^ copies/mL, and 1 × 10^4^ copies/mL. As a negative control, a virus-free dilution was also prepared. Seventy microliters of each recombinant viral dilution was added to UTM and extraction reagent solution.

Supplementary Figure 2. Comparison of Ct values between the Smart Gene SARS-CoV-2 and reference real-time RT-PCR assays. The median difference of Ct values between the Smart Gene SARS-CoV-2 and reference real-time RT-PCR assays was 5.975 (IQR: 3.888-7.777).

Supplementary Table1a. Ct values of nasopharyngeal samples

Supplementary Table1b. Ct values of anterior nasal samples

