## Supplementaly Fig1 for "Clinical Evaluation of the Novel Rapid Nucleic Acid Amplification Point-of-Care Test (Smart Gene SARS-CoV-2) in the analysis of Nasopharyngeal and Anterior Nasal samples"

### Slide 1
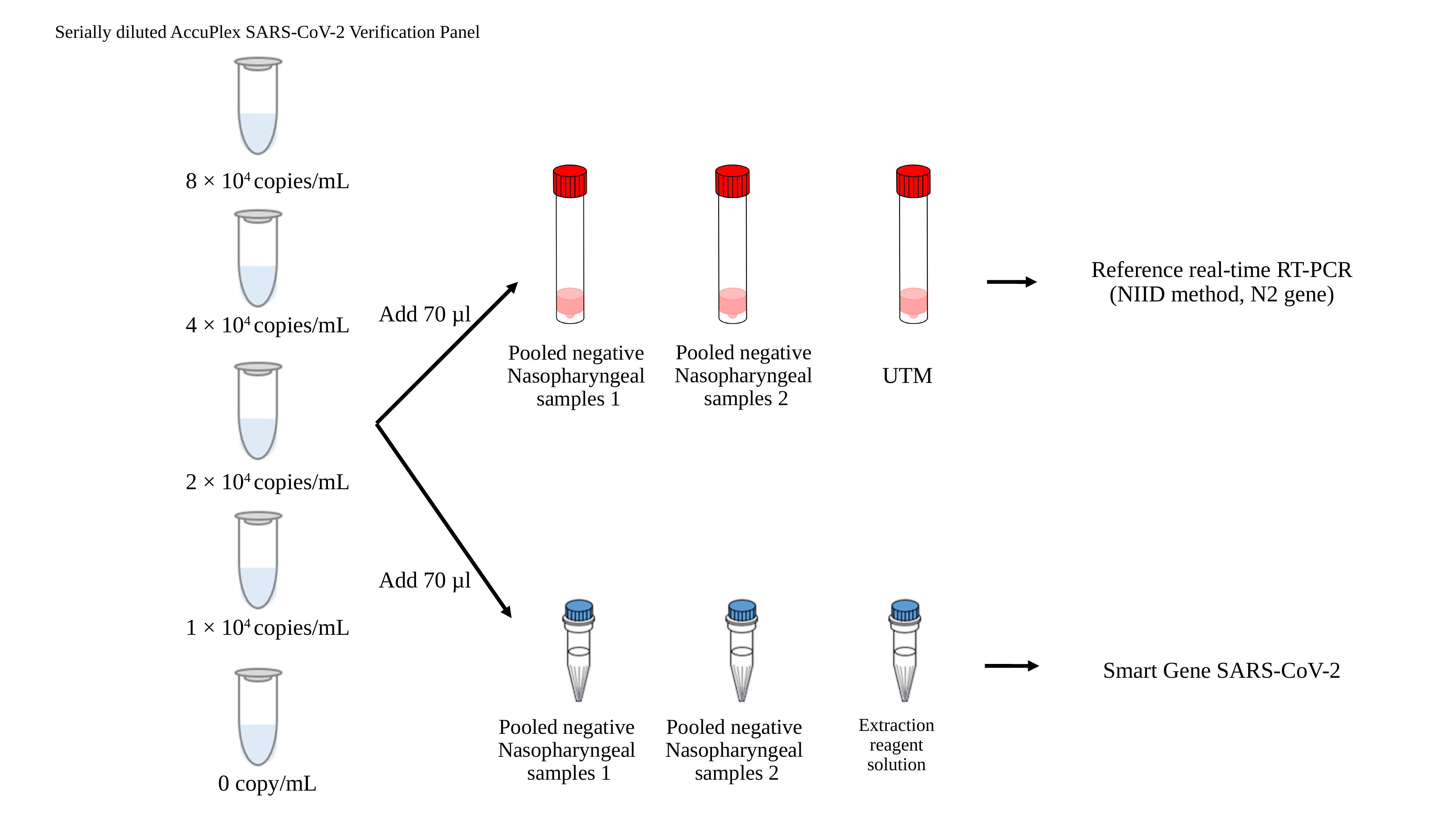

Serially diluted AccuPlex SARS-CoV-2 Verification Panel
8 × 10^4^ copies/mL
Reference real-time RT-PCR
(NIID method, N2 gene)
Add 70 µl
4 × 10^4^ copies/mL
Pooled negative
Nasopharyngeal
samples 2
Pooled negative
Nasopharyngeal
samples 1
UTM
2 × 10^4^ copies/mL
Add 70 µl
1 × 10^4^ copies/mL
Smart Gene SARS-CoV-2
Extraction
 reagent
solution
Pooled negative
Nasopharyngeal
samples 2
Pooled negative
Nasopharyngeal
samples 1
0 copy/mL
