## Supplementary material for "Clinical Evaluation of the Novel Rapid Nucleic Acid Amplification Point-of-Care Test (Smart Gene SARS-CoV-2) in the analysis of Nasopharyngeal and Anterior Nasal samples": Suplementary Fig2

Ct Value

|  | 1 | 2 | 3 | 4 | 5 | 6 | 7 | 8 | 9 | 10 | 11 | 12 | 13 | 14 | 15 | 16 | 17 | 18 | 19 | 20 |
| --- | --- | --- | --- | --- | --- | --- | --- | --- | --- | --- | --- | --- | --- | --- | --- | --- | --- | --- | --- | --- |
| Real-time RT-PCR | 16.56 | 34.69 | 20.87 | 17.51 | 17.23 | 18.69 | 29.9 | 34.36 | 22.33 | 17.49 | 19.65 | 23.28 | 21.68 | 23.07 | 25.12 | 18.91 | 24.51 | 21.22 | 35.26 | 17.77 |
| Smart Gene SARS-CoV-2 | 23 | 40 | 31 | 28 | 25 | 27 | 33 | 38 | 28 | 24 | 23 | 24 | 23 | 37 | 28 | 24 | 37 | 29 | 39 | 24 |
