## Supplementary Table1 for "Clinical Evaluation of the Novel Rapid Nucleic Acid Amplification Point-of-Care Test (Smart Gene SARS-CoV-2) in the analysis of Nasopharyngeal and Anterior Nasal samples"

Supplementary Table1a. Ct values of nasopharyngeal samples

| Sample number | Ct value | |  | Sample number | Ct value | |
| --- | --- | --- | --- | --- | --- | --- |
|  | Reference real-time RT-PCR  (NIID method N2 gene) | Smart Gene SARS-CoV-2 |  |  | Reference real-time RT-PCR  (NIID method N2 gene) | Smart Gene SARS-CoV-2 |
| 1 | 16.6 | 23 |  | 36 | 23.7 | 27 |
| 2 | 34.7 | 40 |  | 37 | 12.3 | 30 |
| 3 | 20.9 | 31 |  | 38 | 34.5 | 39 |
| 4 | 17.5 | 28 |  | 39 | 14.9 | 24 |
| 5 | 17.2 | 25 |  | 40 | 23.5 | 38 |
| 6 | 18.7 | 27 |  | 41 | 22.9 | 25 |
| 7 | 29.9 | 33 |  | 42 | 17.5 | 25 |
| 8 | 34.4 | 38 |  | 43 | 21.7 | 28 |
| 9 | 22.3 | 28 |  | 44 | 20.1 | 34 |
| 10 | 17.5 | 24 |  | 45 | 23.6 | 28 |
| 11 | 19.7 | 23 |  | 46 | 33.4 | 40 |
| 12 | 23.3 | 24 |  | 47 | 19.0 | 25 |
| 13 | 21.7 | 23 |  | 48 | 17.0 | 23 |
| 14 | 23.1 | 37 |  | 49 | 22.2 | 28 |
| 15 | 25.1 | 28 |  | 50 | 18.0 | 23 |
| 16 | 18.9 | 24 |  | 51 | 16.0 | 29 |
| 17 | 24.5 | 37 |  | 52 | 20.5 | 23 |
| 18 | 21.2 | 29 |  | 53 | 17.3 | 23 |
| 19 | 35.3 | 39 |  | 54 | 32.5 | 31 |
| 20 | 17.8 | 24 |  | 55 | 25.7 | 38 |
| 21 | 31.3 | 30 |  | 56 | 29.2 | 40 |
| 22 | 31.3 | 32 |  | 57 | 19.3 | 24 |
| 23 | 19.9 | 26 |  | 58 | 16.9 | 24 |
| 24 | 27.8 | 37 |  | 59 | 17.3 | 24 |
| 25 | 33.9 | 42 |  | 60 | 27.8 | 39 |
| 26 | 33.6 | 38 |  | 61 | 25.1 | 32 |
| 27 | 32.1 | 41 |  | 62 | 17.2 | 23 |
| 28 | 25.0 | 29 |  | 63 | 29.7 | 29 |
| 29 | 17.6 | 24 |  | 64 | 20.1 | 29 |
| 30 | 14.7 | 23 |  | 65 | 27.4 | ND |
| 31 | 19.5 | 28 |  | 66 | 35.3 | ND |
| 32 | 16.5 | 23 |  | 67 | 34.8 | ND |
| 33 | 16.3 | 23 |  | 68 | 36.7 | ND |
| 34 | 24.6 | 31 |  | 69 | ND | 42 |
| 35 | 19.7 | 27 |  | 70 | ND | 33 |

Supplementary Table1b. Ct values of anterior nasal samples

| Sample number | Ct value | |  |  | Ct value | |
| --- | --- | --- | --- | --- | --- | --- |
|  | Reference real-time RT-PCR  (NIID method N2 gene) | Smart Gene SARS-CoV-2 |  | Sample number | Reference real-time RT-PCR  (NIID method N2 gene) | Smart Gene SARS-CoV-2 |
| 1 | 20.2 | 27 |  | 36 | 26.2 | 30 |
| 2 | 22.9 | 27 |  | 37 | 28.9 | 31 |
| 3 | 25.3 | 32 |  | 38 | 27.7 | 27 |
| 4 | 32.0 | 36 |  | 39 | 27.5 | 28 |
| 5 | 36.8 | 41 |  | 40 | 22.9 | 24 |
| 6 | 24.2 | 29 |  | 41 | 27.2 | 30 |
| 7 | 26.1 | 33 |  | 42 | 29.1 | 35 |
| 8 | 25.5 | 30 |  | 43 | 21.0 | 31 |
| 9 | 25.4 | 38 |  | 44 | 31.0 | 34 |
| 10 | 22.0 | 28 |  | 45 | 20.9 | 28 |
| 11 | 26.7 | 31 |  | 46 | 27.6 | 29 |
| 12 | 21.0 | 38 |  | 47 | 29.9 | 37 |
| 13 | 17.0 | 23 |  | 48 | 31.9 | 38 |
| 14 | 18.7 | 23 |  | 49 | 24.0 | 26 |
| 15 | 19.8 | 23 |  | 50 | 34.5 | 38 |
| 16 | 28.6 | 31 |  | 51 | 33.0 | ND |
| 17 | 20.2 | 26 |  | 52 | ND | 38 |
| 18 | 22.9 | 28 |  | 53 | ND | 27 |
| 19 | 34.7 | 40 |  | 54 | ND | 41 |
| 20 | 32.9 | 39 |  |  |  |  |
| 21 | 20.7 | 25 |  |  |  |  |
| 22 | 22.1 | 30 |  |  |  |  |
| 23 | 18.3 | 23 |  |  |  |  |
| 24 | 20.8 | 31 |  |  |  |  |
| 25 | 21.7 | 30 |  |  |  |  |
| 26 | 29.2 | 32 |  |  |  |  |
| 27 | 35.1 | 39 |  |  |  |  |
| 28 | 31.1 | 38 |  |  |  |  |
| 29 | 23.6 | 33 |  |  |  |  |
| 30 | 38.7 | 41 |  |  |  |  |
| 31 | 18.8 | 27 |  |  |  |  |
| 32 | 28.1 | 35 |  |  |  |  |
| 33 | 19.0 | 30 |  |  |  |  |
| 34 | 25.2 | 30 |  |  |  |  |
| 35 | 20.9 | 26 |  |  |  |  |

RT-PCR, reverse transcription polymerase chain reaction; NIID, National Institute of Infectious Diseases; Ct, cycle threshold; ND, not detected.
